# Sooner than you think: a very early affective reaction to the COVID-19 pandemic and quarantine in Argentina

**DOI:** 10.1101/2020.07.31.20166272

**Authors:** F. Torrente, A. Yoris, D.M. Low, P. Lopez, P. Bekinschtein, F. Manes, M. Cetkovich

**Author notes:** first authorship shared (F. Torrente, A. Yoris).

## Abstract

The unique circumstances created by the COVID-19 pandemic pose serious challenges to mood stability and emotional regulation at all ages. Although many people tend to react resiliently to stress, others appear to display emotional anxiety and depression-related symptoms. In this study, we carried out a survey (N = 10,053) during the first week of the general lockdown (quarantine) in Argentina to measure early affective reactions in Argentine adults. Respondents showed substantial anxious and depressive symptoms, with 33 % and 23% of participants reporting possible depressive and anxious syndromes, respectively, with the youngest group (18 to 25 y.o.) showing the highest prevalence of symptoms. Even if prior mental health problems predisposed or aggravated the reaction, participants without prior complaints showed signs of psychological impact. Using linear regression, the most important independent variables related to depressive symptoms was the feeling of loneliness followed by daily stress. In the case of anxious states, the strongest variables were negative repetitive thinking and feeling of loneliness. Other psychological, economic, and social factors are discussed.

This study is in line with previous literature that highlight the importance of the psychological impact of pandemics, but additionally demonstrates that these reactions are present at a large scale immediately after the start of quarantine with very low infectious rates as an early anticipatory adaptive reaction leading to potential negative outcomes from adjustment disorders to major disorders. In addition, the present results provide potentially relevant information about sudden environmental impacts on affective states and specific pathways for anxiety and depression to be expressed. We end by discussing implications for public policy based on considering the most vulnerable groups.

**Key Findings:**

- More than a third of the studied sample (n = 10,053) showed substantial depressive and anxious symptoms between 5 and 7 days after the start of the national quarantine.
- 33.7% of the sample scored above the PHQ-9 cutoff for possible diagnosis of a depressive disorder and 23.2% for an anxiety disorder.
- 28.6% of the participants showed moderate and severe levels of depressive symptoms and 23.2% showed moderate and severe levels of anxiety.
- The youngest participants were affected the most by the situation.
- The most important factor related to depressive symptoms was the feeling of loneliness followed by daily stress.
- The most important factor related to anxious states was negative repetitive thinking followed by feeling of loneliness.
- The existence of previous mental health difficulties aggravated the reactions, but even people who had not sought out treatment previously showed signs of psychological impact.

---

The circumstances created by the COVID-19 pandemic are generating unprecedented challenges to mood stability and emotional regulation of entire populations at once. The fear caused by the threat of a highly contagious disease together with the side-effects of strong prevention measures such as social isolation and quarantine make a powerful environmental combination that could have a significant impact on the mental health of a great number of people.

The spread of fear has been noticed as an important psychological factor in previous recent epidemics such as the 2003 SARS and 2014-2015 Ebola virus epidemics (Taylor, 2019; Shultz et al., 2015). Three common features increase fear spreading: (i) if the infection is new and unpredictable, (ii) its management with isolation, and (iii) a generalized fear of infecting or being infected by others. At the same time, high uncertainty about the future, including the consequences and duration of the pandemic, and continuous presence of alarming news and fatalities contribute to increase fear reactions (Asmundson & Taylor, 2020; Gao et al., 2020). In previous epidemics, the number of patients with mood and anxiety disorders has grown together with the aggravation of symptoms among previously diagnosed patients (Shultz, Baingana, & Neria, 2015; Wu et al., 2009; Taylor, 2019). Furthermore, people affected with psychological disorders may exceed the number of infected cases and would require considerable amounts of additional specialized mental health treatment (Eichelberger, 2007; Taylor, 2019; Normile, 2016; Kluger, 2014; Chang et al., 2004; Person et al., 2004).

At the same time, previous studies have shown that isolation measures during epidemics can cause a significant and lasting psychological impact (Taylor, 2019). Extreme isolation generates a stressful environment accompanied by fears of infection, financial preoccupations, tasks overload, frustration and boredom, inadequate information, insufficient supplies, among other factors. A recent review (Brooks et al., 2020) lists a wide range of psychological manifestations in people under quarantine, such as general psychological symptoms, emotional disorders, depression, low mood, irritability, insomnia, stress symptoms, anger, and emotional exhaustion. If the quarantine is appraised as a negative experience there may be long-term consequences (Brooks et al, 2020). In previous epidemics, a period longer than 10 days of quarantine has been related with a wide range of psychiatric disorders in the general population such as post-traumatic stress disorder (Brooks et al., 2020), anxiety (Taylor, 2019), and depression (Hawryluck et al., 2004). Considering the magnitude of COVID-19 pandemic and the extreme measures undertaken to fight it, it is expected that it can cause at least similar reactions in most populations where the virus has spread rapidly. Recent studies in the earliest affected countries confirm this assumption (Mazza et al., 2020; Qiu et al., 2020; Tian et al., 2020; Wang et al., 2020)

Alongside the medium and long term effects of pandemics and quarantine previously described, it is conceivable that generalized isolation may also provoke a rapid effect on emotional status by different means in at least a subset of the affected population. First, the adoption of strong protective measures such as a quarantine at a national scale may work as a signal of the seriousness of the circumstances that in turn reinforces the magnitude of perceived threat. Second, a strict lockdown means a sudden disruption of everyday routines and social-biological rhythms of millions of people.

Social contact, work activities, physical exercise, sleep, and sexual activity are only some of the areas that may be affected or suppressed during the quarantine. These abrupt changes that convey a reduction in pleasant activities and an increase of aversive experiences may negatively impact on the rate of reinforcement as posited by behavioral theories of depression (Lewinson, 1975; Carvalho & Hopko, 2011) and affect reward processing in more vulnerable individuals through a gene-environment interaction as depicted by more recent neurobiological accounts (Nusslock & Alloy, 2017). In the same vein, irregular social rhythms and circadian rhythm disruption have been associated with a greater risk of developing mood fluctuations and symptomatology in people vulnerable to mood disorders (Ehlers et al., 1988; Russo & Nestler, 2013; Zaki et al., 2018). Third, being isolated may provoke an increase of worries, ruminations, and other kinds of recurrent negative oriented thinking associated with negative affect. This kind of repetitive negative thinking is associated with the development of both depressive and anxious symptoms (Ehering & Watkins, 2008; Drost et al., 2014; Spinhoven et al., 2018; McEvoy et al., 2019). Finally, strong isolation may exacerbate feelings of loneliness in people who were previously experiencing it or even generate new perceived loneliness in certain cases. Social isolation and feelings of loneliness have been linked to a variety of negative mental health outcomes (Cacioppo, et al., 2015; Cacioppo & Cacioppo, 2018).

Meanwhile most studies of previous epidemics did not measure psychological impact in shorter periods than 10 days and were restricted to a limited number of people (Brooks et al., 2020), some recent studies of COVID-19 pandemic report the effects of large-scale quarantines on general population in the earliest stage of isolation measures (Wang et al., 2020; Mazza et al., 2020; González-Sanguino et al., 2020).

Accordingly, the aim of this study was to evaluate the affective impact caused by the COVID-19 pandemic in the first days of the national quarantine ordered by the national authorities in Argentina which banned all small or large social gatherings, outdoor sport activities, closing parks, all non-essential businesses including banks, and restricting public transportation to only essential workers. For this purpose, our study was rapidly conducted after 5-7 days of quarantine. The levels of depression and anxiety in a nationwide sample of the Argentine population were studied using standardized self-report questionnaires together with other related quantitative and qualitative variables that may be related with mood and emotional variations. As a unique element of our sample, the preventive and compulsory social isolation was established in Argentina when there were still few confirmed COVID-19 cases compared to other countries (see Table 1). During the days of the survey the number of cases grew from 301 (or 6.66 per million) to 589 (or 13.03 per million) meanwhile the deaths augmented from 4 (0.09 per million) to 12 (0.27 per million). Because of that, the results of this study may help to understand the very early affective reaction to the pandemic and related preventive strategies.

**Table 1.** Sample of mental health surveys during COVID-19.

| Study | Dates | Country | Sample size | Cases (per million) | Deaths (per million) | Interpersonal contact rank |
| --- | --- | --- | --- | --- | --- | --- |
| Wang et al. (2020) | 01/31 – 02/02 | China | 1210 | 9714 (6.7) – 14399 (10.0) | 213 (0.1) – 304 (0.2) | 32/42 |
| Tian et al. (2020) | 01/31 – 02/02 | China | 1060 | 9714 (6.7) – 14399 (10.0) | 213 (0.1) – 304 (0.2) | 32/42 |
| Gao et al. (2020) | 01/31 – 02/02 | China | 4872 | 9714 (6.7) – 14399 (10.0) | 213 (0.1) – 304 (0.2) | 32/42 |
| Qiu et al. (2020) | 01/31 – 02/10 | China | 52730 | 9714 (6.7) – 40206 (27.9) | 213 (0.1) – 909 (0.6) | 32/42 |
| Huang & Zhao (2020) | 02/03 – 02/17 | China | 7236 | 17211 (12.0) – 70618 (49.1) | 361 (0.3) – 1771 (1.2) | 32/42 |
| Liu et al. (2020) | 02/23 – 04/02 | China | 217 | 77016 (53.5) – 82395 (57.2) | 2445 (1.7) – 3316 (2.3) | 32/42 |
| Bruine de Bruin (2020) | 03/10 – 03/31 | United States | 6666 | 754 (2.3) – 164620 (497.3) | 26 (0.1) – 3170 (9.6) | 13/42 |
| González-Sanguino et al. (2020) | 03/14 – 03/28 | Spain | 3480 | 7641 (163.4) – 83885 (1794.1) | 121 (2.6) – 4858 (103.9) | 8/42 |
| Amerio et al. (2020) | 03/15 – 04/15 | Italy | 131 | 21157 (349.9) – 162488 (2687.4) | 1441 (23.8) – 21069 (348.5) | 11/42 |
| Mazza et al. (2020) | 03/18 – 03/22 | Italy | 2766 | 31506 (521.1) – 53578 (886.1) | 2505 (41.4) – 4827 (79.8) | 11/42 |
| Cui et al. (2020) | 03/20 – 04/01 | China | 892 | 81229 (56.4) – 82295 (57.2) | 3253 (2.3) – 3310 (2.3) | 32/42 |
| Asmundson et al. (2020) | 03/21 – 04/01 | United States | 6854 | 19624 (59.3) – 189618 (572.9) | 260 (0.8) – 4079 (12.3) | 13/42 |
| Our study | 03/24 – 03/27 | Argentina | 10053 | 301 (6.7) – 589 (13.0) | 4 (0.1) – 12 (0.3) | 1/42 |
| Fisher et al. (2020) | 04/03 – 05/02 | Australia | 13829 | 5224 (204.9) – 6767 (265.4) | 23 (0.9) – 93 (3.6) | - |
| Hawke et al. (2020) | 04/08 – 04/29 | Canada | 622 | 17883 (473.8) – 50015 (1325.2) | 380 (10.1) – 2859 (75.8) | 20/42 |
| Chen et al. (2020) | 04/10 – 05/02 | Ecuador | 252 | 4965 (281.4) – 26336 (1492.7) | 272 (15.4) – 1063 (60.2) | - |
| Pierce et al. (2020) | 04/23 – 04/30 | United Kingdom | 17452 | 133495 (1966.5) – 165221 (2433.8) | 21060 (310.2) – 26097 (384.4) | 17/42 |

Results from a Google Scholar search [allintitle:("covid 19"+"mental health"|psychiatry)+survey]] which returned 52 studies and we excluded studies that were unpublished (N=33) or that have imprecise survey dates (N=3). Surveys are ranked by data collection start date. Qiu et al (2020) also included participants from Hong Kong, Macau, and Taiwan and Asmundson et al. (2020) included participants from Canada, but these studies did not specify the proportion of participants from each country surveyed, therefore we report values for the countries with the largest population (China and the United States, respectively). COVID-19 cases and deaths are obtained from ourworldindata.org (source: European CDC). Interpersonal contact rank is how each country ranks regarding their preferred interpersonal distance during conversation with strangers out of 42 countries from Sorokowska (2017).

As another cultural distinctive trait, Argentines may be particularly susceptible to the effects of social isolation given how much they may prefer close contact; one study showed they prefer the closest distance to strangers and acquaintances during conversations out of 42 nationalities (Sorokowska et al, 2017; see Table 1). There are several Argentine close-contact customs that suddenly became dangerous and were disrupted such as the customary cheek kiss when greeting any gender or the custom of drinking a local tea called mate which is generally shared (e.g., at home or at work) using the same cup and straw. We assumed that the combined effect of fear of pandemic and quarantine restrictions would have a significant and rapid impact among Argentines, especially in more vulnerable groups. We hypothesized that the perceived threat, perceived risk of transmission, daily stress, negative repetitive thinking, and feelings of loneliness would be associated with higher affective impact in the first days of the quarantine and we expected younger and older groups to be the most affected.

## Methods

### Participants

This report is based on a sample of 10,053 individuals from Argentina over the age of 18. Gender was reduced to three main categories: female, male, and non-binary. Education profile was segmented into four categories considering the national education system (see Table 2 for details). The family’s basic income was asked in monthly Argentine pesos and converted to three categories (low, medium, and high income). As social media platforms were part of the main delivery system, all participants gave their informed consent asserting to know their privacy would be protected following the Declaration of Helsinki and national laws.

**Table 2.** Sociodemographic data, perception of COVID-19, and attitudes toward quarantine.

|  |  |  |  |  |  |
| --- | --- | --- | --- | --- | --- |
| **Age** | **Mean** | **SD** | **Min** | **Max** |  |
|  | 41.55 | 11.52 | 18 | 84 |  |
| **Gender** | **Female** | **Male** | **N-B1** | **NA2** |  |
| (% of the sample) | 83.4 | 16.3 | 0.1 | 0.1 |  |
| **Education (years)** | **7-12** | **12-15** | **15-20** | **20+** |  |
| (% of the sample) | 5.5 | 14.4 | 34.6 | 45.4 |  |
| **Family income** | **Low** | **Medium** | **High** |  |  |
| (% of the sample) | 42.3 | 39.5 | 18.2 |  |  |
| **Perceived threat of COVID-19** | **Total Sample** | **18-25** | **26-44** | **45-64** | **65+** |
| (% of the sample) |  |  |  |  |  |
| Low | 8.2 | 19.7 | 8.5 | 5.0 | 1.9 |
| Medium | 28.4 | 43.4 | 30.6 | 21.8 | 13.9 |
| High | 45.2 | 31 | 45.3 | 48.4 | 49.6 |
| Very high | 18.2 | 6 | 15.6 | 24.7 | 34.6 |
| **Perceived risk of transmission** | **Total Sample** | **18-25** | **26-44** | **45-64** | **65+** |
| (% of the sample) |  |  |  |  |  |
| Low | 14.1 | 24.6 | 13.2 | 13.2 | 11.3 |
| Medium | 36.9 | 40.9 | 36.7 | 36.7 | 31.2 |
| High | 38.3 | 29.1 | 39.0 | 38.8 | 44.4 |
| Very high | 10.7 | 5.5 | 11.1 | 11.2 | 13.2 |
| **Agreement with quarantine** | **Total Sample** | **18-25** | **26-44** | **45-64** | **65+** |
| (% of the sample) |  |  |  |  |  |
| Low | 0.6 | 1.1 | 0.5 | 0.4 | 1.3 |
| Medium | 1.0 | 1.1 | 1.2 | 0.6 | 0.3 |
| High | 3.5 | 4.6 | 3.7 | 3.0 | 2.2 |
| Very high | 95.0 | 93.2 | 94.6 | 95.9 | 96.2 |
| **Perceived efficacy of quarantine** | **Total Sample** | **18-25** | **26-44** | **45-64** | **65+** |
| (% of the sample) |  |  |  |  |  |
| Low | 0.5 | 1.0 | 0.4 | 0.4 | 1.3 |
| Medium | 1.1 | 2.2 | 1.2 | 0.8 | 0.3 |
| High | 5.6 | 8.8 | 5.7 | 4.8 | 4.1 |
| Very high | 92.8 | 88.0 | 92.7 | 94.0 | 94.4 |
| **Adherence to quarantine** | **Total Sample** | **18-25** | **26-44** | **45-64** | **65+** |
| (% of the sample) |  |  |  |  |  |
| Yes | 94.5 | 95.9 | 94.1 | 94.8 | 97.8 |
| No | 2.1 | 0.7 | 2.2 | 2.3 | 0.3 |
| Partial | 3.4 | 3.4 | 3.7 | 3 | 1.9 |

### Instruments

A survey was designed to evaluate different variables associated with the psychological impact of the pandemic and quarantine. The survey included two standardized questionnaires to assess the severity of symptoms of the depressive and anxious series:

### Patient Health Questionnaire-9 (PHQ-9)

The PHQ-9 is a brief self-report scale composed of nine items based on the DSM-IV criteria for the diagnosis of major depressive episode. It has been developed to assess the presence and severity of depressive symptoms in primary care and in the community and to establish a tentative diagnosis of a depressive episode. The Argentine version of PHQ-9 (Urtasun et al., 2019) had high internal consistency (Cronbach’s alpha = 0.87) and satisfactory convergent validity with the BDI-II scale [Pearson’s r= 0.88 (p < 0.01)]. The cut-off points established by Urtasun et al. (2019) were used to evaluate the possible diagnosis of depression and the ranges of severity in the present study. A score of 8 or more indicated a possible diagnosis of major depression according to DSM-IV. The cut off points for severity ranges were 6–8 for mild cases, 9–14 for moderate, and 15 or more for severe depressive symptoms respectively.

### Generalized anxiety disorder-7 (GAD-7)

The GAD-7 is a brief 7-item self-report questionnaire designed to identify probable cases of generalized anxiety disorder and to assess the severity of symptoms. GAD-7 also proved to have good sensitivity and specificity as a screener for panic, social anxiety, and posttraumatic stress disorder (Kroenke et al. 2008). The original version of the questionnaire (Spitzer et al., 2006) showed very high internal consistency (Cronbach’s alpha = 0.92) and satisfactory convergent validity with the Beck Anxiety scale [r= 0.72(p < 0.01)] and with the anxiety subscale of the Symptom Checklist-90 (r = 0.74). The Spanish version for Argentina of the GAD-7 was used in this case (downloaded from: https://www.phqscreeners.com/select-screener). The GAD-7 was utilized in previous studies in Argentina (Gargoloff et al., 2016) and showed a high internal consistency for the sample of the present study (Cronbach’s alpha = 0.90). To establish the severity levels of the current sample, the cut-off points were used according to Spitzer et al. (2006). Accordingly, scores of 5, 10, and 15 were taken as the cut-off points for mild, moderate and severe anxiety, respectively. A score of 10 or greater was considered as indicative of the presence of a possible anxiety disorder.

### Sociodemographic characteristics

Potentially relevant general characteristics such as age, gender, family income, and level of education were surveyed. Furthermore, being in current treatment for a previous mental health condition was asked as a proxy for a potential pre-existing disorder.

### COVID-19’s perception and attitudes towards quarantine

The survey also included questions created *ad hoc* to evaluate variables related to the pandemic and quarantine. Perception of threat of COVID-19 and perception of the risk of transmission were explored as single dimensions in a scale from 0 (“not at all”) to 10 (“extreme”).

Attitudes toward quarantine were evaluated considering three self-reported dimensions: adherence to the measure (“yes, no, partially”), agreement with the norm (from 0 “not at all” to 10 “completely agree”), and trust about its effectiveness as a preventive health tool (from 0 “not at all effective” to 10 “very effective”).

### Daily stress

Impact in daily life was assessed within five domains: work, household chores, physical exercise, leisure, activities with children, and relationship with other adults. For each of the areas, the participants had to rate how difficult it was for them to carry out the daily activities compared to the moments prior to quarantine (from very difficult to very easy). A general index of daily stress was calculated summing up the scores of each of the six domains assessed (with a score of −2 for “very difficult”, −1 for “difficult”, 0 for “neutral”, 1 for “easy”, and 2 for “very easy”). As a result, the index varies from −12 (more negative daily stress scenario) to 12 (more positive daily stress scenario).

### Feelings of loneliness

Loneliness was measured as a single dimension asking participants to report how lonely they feel from 0 (‘Not at all’) to 10 (‘Extremely’) in the last week.

### Negative repetitive thinking

As stated by Ehring and Watkins (2008) individuals with emotional disorders usually report excessive and repetitive thinking about their current concerns, problems, past experiences, or worries about the future. In the current study, we explored this dimension by assessing the presence of an increased number of negative thoughts related with past, future, or interpersonal concerns since the beginning of the quarantine. For each of these options there was a categorical (yes/no) answer. Negative repetitive thinking was considered present when at least one of the options was selected.

### Procedures

The survey was distributed through different social media networks (Facebook, Twitter, Instagram, WhatsApp) and through email. The questionnaire was enabled on 03/24/2020 and the recruitment of the present sample was completed in 32 hours. The official start of the national quarantine in Argentina was established at 0:00 on 03/20/20, so the responses obtained correspond to a period of between 5 and 7 days of isolation.

### Statistical Analysis

Comparisons between groups were made using one-way ANOVA followed by Tukey’s HSD or Tamhane 2 for post hoc comparisons when appropriate. Correlations between measures were carried out by using the Pearson correlation coefficient with Bonferroni correction for multiple comparisons using α = 0.05. When analyzing categorical variables, the Pearson chi-square test was used. Multiple linear regression was employed to develop explanatory models for the two main dependent variables (depression and anxiety).

## Results

### Sociodemographic characteristics

The mean age of participants was 44.55 years (SD= 11.52; min= 18, max= 84). Female gender was the most representative (83.4 %) and few cases reported a non-binary gender identification or preferred not to answer (0.3 %). There were participants from all of the country’s provinces. The sample was well distributed across income levels. Regarding education, even if all ranks were represented, there was a tendency towards over-representing the higher levels (see details in Table 2). A smaller proportion of the participants (15.8%) stated that they were under psychological or psychiatric treatment for a previous mental health condition.

### Perception of COVID-19 and attitudes toward the quarantine

Regarding COVID-19’s threat perception (see Table 2), most participants selected high (45.2 %) and medium (28.4 %) ratings, while very high (18.2%) and low (8.2 %) were the less frequent responses. When segmented by ages, an opposite pattern was found between youngest and oldest participants. Perception of COVID-19 threat was rated as very low by 19.7 % of participants of 18-25 range, meanwhile only 6% of them perceived a very high threat. In contrast, 34.6% of elders (65+) perceived threat of COVID-19 as very high, and only 1.9% estimated a very low threat.

In the case of perceived risk of transmission, medium (36.9 %) and high (38.3 %) ratings were the most frequent responses for the whole sample, followed by very low (14.1%), and very high risk (10.7 %). Segmented by age, risk of transmission results presented a similar trend than perceived risk, but with milder differences (see Table 2).

Attitudes towards quarantine in the early stage of confinement were highly positive across the three dimensions for all age groups (see Table 2). Most participants perceived themselves as compliant with the quarantine (94.5%), most agreed with its implementation (98.5% selected high and very high ratings), and most perceived quarantine as an efficacious intervention for controlling pandemics (98.4% with high and very high ratings).

### Daily life stress

Ratings of subjective difficulty while doing daily life activities during quarantine showed that 49% of participants find difficult or very difficult to do work duties, followed by exercise (48.1 %) and leisure activities (31.8%) as the more difficult tasks to perform. Household chores, activities with children and relationships with other adults were perceived as less difficult (Table 3).

**Table 3.** Daily life impact.

| **Perceived difficulty**<br>**of activities during**<br>**quarantine**<br>**(total sample %)** | **Easy** | **Very easy** | **Neutral** | **Difficult** | **Very difficult** |
| --- | --- | --- | --- | --- | --- |
| Work | 12.7 | 9.7 | 28.6 | 24.4 | 24.6 |
| Physical activity | 18.2 | 5.9 | 27.7 | 30.4 | 17.7 |
| Household chores | 39.5 | 25.3 | 26.5 | 6.8 | 1.8 |
| Relationship with<br>others (adults) | 29.5 | 19.7 | 28.2 | 14 | 8.6 |
| Activities with<br>children | 25 | 38.8 | 23.7 | 6.8 | 5.8 |
| Leisure activities | 26.9 | 12.2 | 23.5 | 19.6 | 12.2 |
| **Stress when**<br>**leaving home** | **Very low** | **Low** | **Medium** | **High** | **Extreme** |
| Total sample % | 25.0 | 10.9 | 20.4 | 23.9 | 19.7 |

Considering stress when leaving home, 43.6 % of the total sample reported high and extreme levels of stress, followed by a 20.4 % that rated medium, 10.9 % low and 25 % very low stress.

### Psychological impact

#### Depressive symptoms

PHQ-9’s score analysis revealed that 53.0 % of responders did not show depressive symptoms, followed by 18.5 % that showed mild, 18.1 % moderate and 10.5 % severe symptoms (Table 4). Moderate and severe symptoms together reached 28.6 % of the total sample. Divided into age subgroups, the 18-25 group was the most depressed with 52.4 % of participants reporting moderate and severe scores, followed by 25-44 with 29.5 %, then 45-64 with 21.9 and 65+ with 15.1 %. Considering the cutoff score of the PHQ-9 in Argentina (>8), it was found that a 33.7% of the sample reached the level for the possible diagnosis of a major depressive episode. In contrast, when analyzing the number of symptoms required for the diagnosis according to DSM-V, an 8.8% of participants met the criteria of 5 or more symptoms, including depressed mood or loss of interest. Regarding specific symptoms, 8.3% of the sample reported suicidal thoughts (item 9), meanwhile the most frequent symptoms were poor appetite or overeating (item 5 reported by 31.3% of the sample) and trouble falling or staying asleep, or sleeping too much (item 3 reported by 25.4% of the sample).

**Table 4.** Affective impact.

|  | **Total sample** | **18-25** | **25-44** | **45-64** | **65+** |
| --- | --- | --- | --- | --- | --- |
| **Depression scores (PHQ-9)** |  |  |  |  |  |
| (M/SD) | 6.53 (5.63) | 9.76 (6.23) | 6.72 (5.57) | 5.54 (5.24) | 4.50 (5.10) |
| **Depression severity (PHQ-9)** |  |  |  |  |  |
| (% of the sample) |  |  |  |  |  |
| None | 53.0 | 28.5 | 51.2 | 61.0 | 68.7 |
| Mild | 18.5 | 19.0 | 19.3 | 17.1 | 16.3 |
| Moderate | 18.1 | 30.0 | 18.9 | 14.4 | 8.2 |
| Severe | 10.5 | 22.4 | 10.6 | 7.5 | 6.9 |
| **Depression diagnosis (PHQ-9)** |  |  |  |  |  |
| (% of the sample) |  |  |  |  |  |
| Not Depressed | 66.3 | 42.2 | 65.1 | 73.4 | 79.6 |
| Possible Depressed | 33.7 | 57.8 | 34.9 | 26.6 | 20.4 |
| **Anxiety scores (GAD-7)** |  |  |  |  |  |
| (M/SD) | 6.32 (5.27) | 7.69 (5.50) | 6.67 (5.37) | 5.51 (4.93) | 4.55 (4.49) |
| **Anxiety Severity (GAD-7)** |  |  |  |  |  |
| (% of the sample) |  |  |  |  |  |
| None | 45.1 | 35.6 | 42.0 | 51.8 | 62.1 |
| Mild | 31.6 | 29.6 | 32.9 | 30.6 | 24.8 |
| Moderate | 13.6 | 20.9 | 14.6 | 10.5 | 8.2 |
| Severe | 9.6 | 13.9 | 10.6 | 7.1 | 5.0 |
| **Anxiety diagnosis (GAD-7)** |  |  |  |  |  |
| (% of the sample) |  |  |  |  |  |
| Not clinically anxious | 76.8 | 65.2 | 74.8 | 82.4 | 86.8 |
| Possible clinical anxiety | 23.2 | 34.8 | 25.2 | 17.6 | 13.2 |
| **Feelings of Loneliness** |  |  |  |  |  |
| (% of the sample) |  |  |  |  |  |
| Low | 65.4 | 51.3 | 66.0 | 67.9 | 66.8 |
| Medium | 14.4 | 19.9 | 14.2 | 13.5 | 12.5 |
| High | 13.8 | 20.1 | 13.7 | 12.5 | 11.9 |
| Extreme | 6.3 | 8.7 | 6.0 | 6.0 | 8.8 |

Between groups comparison (one-way ANOVA) showed that participants in current mental health treatment had significantly higher levels of depression (*F*(1.10048) = 221.30, *p* < .001, |^2^ = .022). However, participants without current treatment also exhibited elevated rates of depressive symptomatology, since 31.4% of them scored above the cut-off for a possible diagnosis of depression and 26.2% rated moderate and severe levels of depressive symptomatology.

There were also differences in depression scores between age groups (*F*(3,10049)= 142.65, *p* < .001, |^2^ = .041). Post-hoc comparisons showed differences between the four groups (18-25 > 26-45 > 46-64 > 65+). As well, female participants were more depressed than males (*F*(1,10021) = 80.99, *p* < .001, |^2^ = .008). Finally significant differences in depression scores between income groups were found (*F*(2,10047) = 67.58, *p* < .001, |^2^ = .013). Post-hoc comparisons revealed differences between the three groups (low > medium > high; *p* < .001).

#### Anxiety severity

GAD-7’s ratings revealed that 45.1 % of participants showed no anxiety symptoms, 31.6 presented mild symptoms, followed by a 38.4 % with moderate and an 8.1 % with severe symptoms (Table 4). Divided in age subgroups, the 18-25 range was the most anxious (34.8 % with moderate or severe symptoms), followed by 25-44 (25.2 %), 45-64 (17.6 %) and 65+ (13.2 %). Regarding the cut-off for a possible anxiety disorder, 23.2% of participants showed scores of 10 or greater, suggesting the need for further evaluation. As in the case of depression, being on current treatment for a mental health condition (*F*(1,10048) = 229.072, *p* < .001, |^2^ = .022) and being a woman (*F*(1,10021) = 103.91, *p* < .001, |^2^ = .010) were associated with higher levels of anxiety. Comparison between income groups revealed significant differences in anxious scores (*F*(2,10047) = 14.42, *p* < .001, |^2^ = .003). Post-hoc comparisons revealed differences between the three groups (low > medium = high; *p* < .001).

#### Feeling of loneliness

65.4 % of the total sample did not feel lonely during the first days of the quarantine, while a 14.4% rated medium, 13.8 % high and 6.3% extreme loneliness feelings. More intense feelings of loneliness appeared in groups of 18-25 (28.8 % in high and extreme ratings) and 65+ (20.7 %).

#### Negative repetitive thinking

65.3% of the total sample expressed at least one kind of negative repetitive thinking during the first days of quarantine. This group had significantly more depressive (*F*(1,10051) = 1294.045, *p* < .001, |^2^ = .114) and anxiety symptoms (*F*(1,10051) = 1677.678, *p* < .001, |^2^ = .143) than the group without negative repetitive thinking.

### Association between psychological variables

Correlation analysis found several significant associations between relevant variables (Table 5). Age was negatively associated with depression (r = −.199, p < .01). Loneliness feelings were positively associated with both depression and anxiety (r =.424, p < .01 and r = .357, p < .01 respectively). Daily stress index was negatively correlated with depression (r = −.332, p < .01) and anxiety (r = −.305, p < .01), being a more negative index a signal of higher stress. Perceived threat was positively associated with age (r = .265, p < .01) and perceived risk of transmission (r = .467, p < .01).

**Table 5.**
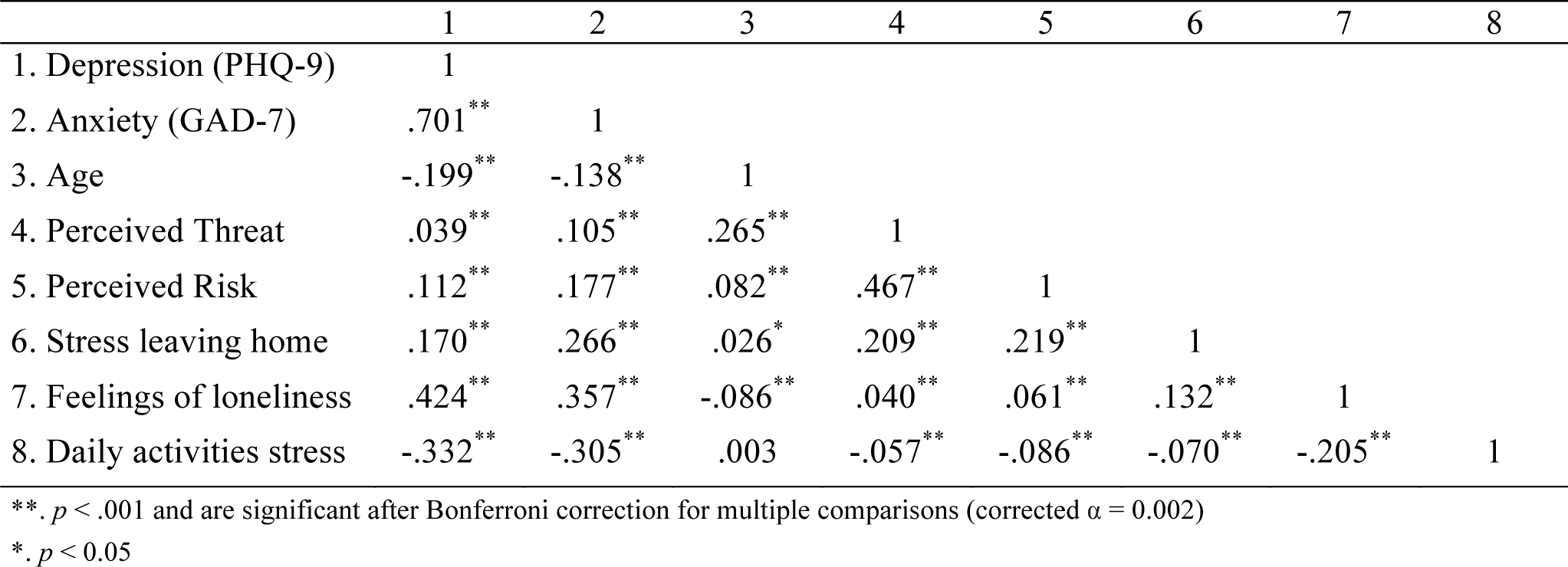
Correlations.

|  | 1 | 2 | 3 | 4 | 5 | 6 | 7 | 8 |
| --- | --- | --- | --- | --- | --- | --- | --- | --- |
| 1. Depression (PHQ-9) | 1 |  |  |  |  |  |  |  |
| 2. Anxiety (GAD-7) | .701** | 1 |  |  |  |  |  |  |
| 3. Age | -.199** | -.138** | 1 |  |  |  |  |  |
| 4. Perceived Threat | .039** | .105** | .265** | 1 |  |  |  |  |
| 5. Perceived Risk | .112** | .177** | .082** | .467** | 1 |  |  |  |
| 6. Stress leaving home | .170** | .266** | .026* | .209** | .219** | 1 |  |  |
| 7. Feelings of loneliness | .424** | .357** | -.086** | .040** | .061** | .132** | 1 |  |
| 8. Daily activities stress | -.332** | -.305** | .003 | -.057** | -.086** | -.070** | -.205** | 1 |

\*\* . $p < .001$ and are significant after Bonferroni correction for multiple comparisons (corrected $\alpha = 0.002$ )
\*. $p < 0.05$

Perceived risk of transmission was also correlated with anxiety (r = .177; p < .01). Finally, stress when leaving home was correlated with anxiety (r = .266, p < .01), perceived threat (r = .209, p < .01), and perceived risk of transmission (r = .219, p <.01).

### Explanatory models of depression and anxiety

A multiple linear regression analysis was carried to evaluate the explanatory role of the different variables over depression and anxiety. A significant regression model for depression was found (F = 610.229, p < .01, R^2^ =.327) with feeling of loneliness, daily stress, negative repetitive thinking, age, gender, current mental health treatment, perceived risk of transmission, and family income as significant independent variables. The three main variables (feeling of loneliness, daily stress, and negative repetitive thinking) explained 28.8% of variance in depressive symptoms, meanwhile the rest of the variables explained the remaining 3.9%.

In the case of anxiety, a significant regression model was found (F = 535.288, p < .01, R^2^ =.298) with negative repetitive thinking, feeling of loneliness, daily stress, perceived risk, age, gender, current mental health treatment, and perceived threat as variables. The three main variables (negative repetitive thinking, feeling of loneliness, and daily stress) explained 26% of variance in depressive symptoms, meanwhile the rest of the variables explained the remaining 3.8%.

## Discussion

This study aimed to quickly measure the early affective impact of the COVID-19 pandemic and of the mitigation strategies based on social isolation. Despite only having spent a few days in quarantine, a noticeable impact on the mood and anxiety of the participants was observed. A striking 33.7% of the sample overpassed the cut-off criteria for the possible diagnosis of major depression recently reported in Argentina (Urtasun et al., 2019) and a 23.7% of participants scored above the cut-off for clinical anxiety (Spitzer et al., 2006). These figures are larger than expected according to previous epidemiological studies in Argentina. A recent national representative community survey found a twelve-month prevalence of 5.7% for any mood disorder and of 9.4% for any anxiety disorder (Stagnaro et al., 2018). Another study in two cities of Argentina employing the PHQ-9 found a prevalence of major depressive episodes of 5.6% (4.2% for men and 7.0% for women) and 9.5% (5.1 for men and 13.6% for women) respectively (Daray et al., 2017). Despite negative emotionality was usually reported in previous epidemics and pandemics (Shultz, Baingana, & Neria, 2015; Taylor, 2019), the severity in the first stages of the confinement and the scale of the impact in large populations should be considered a novelty. Furthermore, our results confirm the findings reported in different cultural contexts affected by the COVID-19 pandemic (Wang et al., 2020; Mazza et al., 2020; González-Sanguino et al., 2020) but this study is unique in that there were very few COVID-19 cases during the survey (301 or 6.66 per million – 589 or 13.03 per million). We can interpret these findings as an indication of a very early anticipatory fear reaction and as the consequence of the sudden disruption of normalcy provoked by the start of a very strict and national-wide quarantine.

However, a note should be made about the interpretation of self-reported scores of depression and anxiety in the context of the present study. It is recognized that self-report methods may overestimate the rate of psychiatric disorders in comparison with the more reliable gold standard of diagnostic interviews. A recent meta-analysis about the use of the PHQ-9 for the screening of major depressive episodes in primary care found that approximately half of patients with positive screens could be false positives (Levis et al., 2019). In fact, in the current study when we considered the number of symptoms required for the diagnosis of major depression instead of the cut-off score, the rate of participants that met the criteria decreased to an 8.8%. Therefore, it is highly probable that many of the states reported by affected participants would correspond to adjustment disorders or other transitory manifestations of troubled affect, instead of a major depressive disorder or an anxiety disorder. According to DSM-V (APA, 2013), the presence of emotional or behavioral symptoms in response to an identifiable stressor is the essential feature of adjustment disorders. Stressors may affect large groups or entire communities, as is the case of COVID-19, and the onset of symptoms may appear within a few days of the occurrence of the stressor, as revealed in the current study. The duration of distress is relatively brief (a few months) but if the stressor or its consequences persist, the adjustment disorder may adopt a more persistent form (APA, 2013). Hence, the main findings of this survey and other similar investigations worldwide could be considered in a dimensional frame. From this perspective, a big environmental stressor with an abrupt beginning such as the COVID-19 pandemic and subsequent quarantine may have an immediate effect over affective states of the population in a continuum that goes from normative stress reactions to anxiety disorders or major depression, going through intermediate and transitory forms such as adjustment disorders. Thus, the observed results of anxiety and depressive symptomatology could be considered the overall emotional cost of the forced and fast adaptation to the new hazardous circumstances.

Another important finding was that the youngest participants were affected most, with an inverse relationship between age and emotional symptoms. We can venture different hypotheses to explain this result. On the one hand, young people may feel more limited in their active social life and show more blatant need for contact and physical activity than older age groups. The sudden restriction may have implied a relatively bigger change on the lifestyle and routines for younger people. On the other hand, since they have a lower perception of threat and feel less susceptible to COVID-19 than the other groups, it is possible that the cost-benefit ratio of the measures were perceived as more disadvantageous for them. The distance between perceived risk and the preventive measures taken may produce a sound cognitive dissonance. In our sample, however, the rate of self-reported adherence to such measures was very high for young participants. Hence, we can hypothesize that compliance occurred at expense of a high emotional cost. Simultaneously, this high adherence among young adults may have been caused by the fear of transmitting COVID-19 to older populations, which could – along with possible economy related anxiety and domestic-stress related stress– explain higher levels of anxiety observed in younger adults. Conversely, since older people feel more vulnerable, they may have strong personal motivations to accept restrictions and therefore they could assimilate them at a lower emotional cost. Furthermore, we can assume that the group of older adults who completed the online survey is an active group connected to others through technology, which could operate in some cases as a protective factor for loneliness (Nowland, Necka & Cacioppo, 2018). This would especially apply to the +65 group that showed a lower level of impact than expected.

Among the activities most affected by quarantine were work and physical activity. Difficulties in accommodating work to exceptional circumstances could have operated as a stressor that worsened people’s psychological adjustment to quarantine. For its part, physical activity is a powerful stimulant that exerts a positive and sustained effect on mood, as witnessed by its use for the treatment of depression (Biddle, 2016). Therefore, we can speculate that restrictions on physical activity may have contributed to the observed effects on mood and anxiety. Again, the lack of physical activity may have had a stronger negative impact in younger people.

Regarding gender differences, women showed significantly higher depression scores than men. Even though it is a well-known fact that women have an increased incidence of depression and anxiety disorders (see Stagnaro et al., 2018 for gender differences in prevalence rates in Argentina), contextual factors could interact in the present circumstances. Daily stress may have been incremented for many women in charge of household chores and child care in parallel with home-based work (Unicef, 2020).

Another component associated with depression and anxiety was the presence of a previous treatment for mental health problems. As noted in other studies related to COVID-19, a pre-existing mood or anxiety disorder is associated with increased levels of psychological distress (Asmundson et al., 2020; van Roekel et al., 2020). However, in the current study the psychological impact was also substantial in many of the participants who did not report being in treatment for a previous condition. Therefore, the existence of pre-existing psychological or psychiatric difficulties could act as an aggravating factor, but it does not explain the impact observed in the total sample. Similarly, participants belonging to a low-income group appeared to be more vulnerable to the impact of the stressor.

From a theoretical point of view, the obtained pattern of results seems to support social and behavioral theories of depression. The drastic reduction of social contact may have increased negative feelings and negative emotionality (Joiner, 1999; Lewinsohn, Gotlib, & Seeley, 1997). Congruently, feelings of loneliness appeared as the most important factor associated with emotional symptoms as evidenced in the multiple regression model, above other psychological variables such as the fear of transmission or the symptoms attributed to the disease. This finding confirms the importance of this social element in the regulation of mood as different studies have been showing worldwide (Cacioppo & Cacioppo, 2018; Nowland, Necka, & Cacioppo, 2018; van Roekel et al., 2018). Moreover, the circumstances created by the pandemic conveyed a considerable decrease of rewarding experiences and provoked a general reduction of behavioral activity in the population which also have been associated to emotional problems (Carvalho & Hopko, 2011; Kasch, Rottenberg, Arnow, & Gotlib, 2002). Sadness and depression are common responses to loss and negative life events. High levels of depressive symptoms may result from the experience of concrete losses such as social proximity, gathering, fitness activities, reduced income, and interrupted routines in general, among other reduced sources of reward. As shown by classic behavioral models (Lewinson, 1975), the COVID-19 pandemic as a salient environmental stressor disturbed normal behavioral patterns and routines, which in turn altered the balance between rewards and punishments, provoked social isolation, increased self-focus and negative repetitive thinking, and then was expressed as negative affect. Further worsening or maintenance of negative affective reactions will depend on the persistence of the stressors and on the co-occurrence of additional risk factors such as pre-existent mental health disorders or other socio-economic vulnerabilities.

Regarding anxiety symptoms, the concept of health anxiety has been invoked as a potentially useful framework for conceptualizing reactions to pandemics (Asmundson & Taylor, 2020). A large-scale textual analysis of posts from mental health forums on Reddit showed a significant increase in health anxiety topics during COVID-19 and showed support groups for different disorders becoming more linguistically similar to the support group for health anxiety (Low et al., 2020). Health anxiety refers to the tendency to become alarmed by illness related stimuli. People with excessively high levels of health anxiety, compared to less anxious people, are more afraid about perceived health threats and more concerned with the likelihood and seriousness of becoming ill (Taylor, 2019). In agreement with this view, we found that levels of anxiety, perceived susceptibility to transmission, perceived threat, and stress when leaving home were intertwined variables. Also, our study confirms the central role that repetitive negative thinking plays in explaining anxiety as well as depressive symptoms (Drost et al., 2014; Spinhoven et al., 2018). Notably alongside repetitive negative thinking, feelings of loneliness, and daily stress also explained depressive and anxiety manifestations, suggesting shared transdiagnostic roots between the two target conditions (Dalgleish et al., 2020; Krueger & Eaton, 2015).

The present study has several limitations. First, the survey was disseminated incidentally. Nevertheless, all of the country’s regions were sampled and the most important urban conglomerates where 80% of Argentine population lives represented 87% of the sample. Second, as noted previously, self-report measures have limited reliability for making diagnosis. It is important to prudentially consider these present results and to avoid jumping to clinical conclusions. Complementary and more precise procedures should be adopted to confirm or reject any assumption of diagnosis. Third, our sample was unbalanced in gender with female being overrepresented over other options. As we mentioned before, female gender is associated with increased rates of anxiety and depression in epidemiological studies, so sampling bias may have inflated the global figures of our whole study. Fourth, while we intended to measure early emotional reactions, further assessments should establish if the observed impact remains, increases, or decreases during longer periods of time. Fifth, due to the observational nature of the study, it is not possible to disentangle the effects produced by the pandemic itself from the impact of the quarantine, although COVID-19 cases were low during this survey. Thus the results should be interpreted as the consequences of the combination of the two factors.

Against these limitations, a series of strengths of the study should be mentioned: the large sample size, a short-time reaction survey ran within 32 hours just 5 days after the start of a nationally implemented rigorous quarantine in a country with low infection rate, a considerable representativeness at the national level, an adequate distribution according to family income, and the use of standardized instruments for measuring the main outcomes.

In closing, we can assert that the psychological impact observed in the present study due to the COVID-19 pandemic and quarantine may have policy implications. To mitigate the consequences of the current circumstances and of future similar events, different actions would be needed: 1) early identification and monitoring of people and groups at risk of incidents related with their mental health; 2) implementation of multi-level interventions for the identified persons and groups; 3) interventions aimed at connecting and supporting isolated people who may experience strong feelings of loneliness such as volunteer systems for the elderly; 4) recommendations and measures to mitigate the stress generated by abnormal work circumstances with a gender perspective; 5) strengthen communication and interventions directed to reduce the emotional costs of the younger population. Regarding mental health interventions, it will be important to explore the use of combined transdiagnostic treatments given the general rise in anxiety and depressive symptoms (Newby et al., 2015; Martin et al., 2018; Dalgleish et al., 2020).

In conclusion, our findings indicate that we can expect a rapid affective reaction at population scale when a large environmental stressor invokes a generalized increase of health anxiety and profoundly disturbs daily routines and social dynamics. Since the pandemic suddenly altered the pre-existent sense of normalcy of an entire population, a big affective shock seems to be the most natural adaptive reaction.

## Data Availability

Only the data published in the document is available for publication.

## Author Contribution Statement

F.T. and A.Y. conceived the design, performed data analysis, and drafted the manuscript. D.M.L. contributed to data analysis and writing. All authors provided feedback on design, analyses and reviewed the manuscript.

